# The impact of prolonged walking on fasting plasma glucose in type 2 diabetes: A Randomised controlled crossover study

**DOI:** 10.1101/2023.02.20.23286165

**Authors:** Anxious J. Niwaha, Lauren R. Rodgers, Andrew T. Hattersley, Robert C Andrews, Beverley M. Shields, Moffat J. Nyirenda, Angus G. Jones

**Author notes:** Corresponding author Angus Jones, (Institute of Biomedical and Clinical Science, College of Medicine and Health, University of Exeter Medical School, EX2 5DW, Exeter, UK). Joint last authors (Moffat J. Nyirenda, Angus G. Jones). Disclosure Statement: The authors have nothing to disclose. Pan African Clinical Trial Registry (https://pactr.samrc.ac.za/) (PACTR202009486614518).

## Abstract

**Aims:** In many low-income countries, fasting glucose is the primary measure of glycaemic control used for treatment titration, as HbA1c is often unavailable or unaffordable. Many patients in these countries walk long distances to the clinic, but the impact of walking on fasting glucose in type 2 diabetes is unknown. We aimed to determine whether this prolonged walking affects the reliability of fasting plasma glucose as a measure of glycaemic control.

**Methods:** In a randomised crossover trial, the change in glucose from baseline in the fasting state was compared between walking on a treadmill at a predetermined speed of 4.5 km/hour for 1 hour and not walking (resting) in people with type 2 diabetes. The pre-specified primary outcome was glucose at 1 and 2 hours.

**Results:** 45 participants were enrolled and all completed both visits. 21/45 (46.7%) were female, and the median age was 51. Glucose during and after walking was similar to glucose while at rest; glucose difference (walking minus rest) was -0.15 (95% CI: -0.55, 0.26) and -0.10 (95% CI: - 0.50, 0.31) mmol/L at 1 and 2 hours respectively, p>0.4 for both).

**Conclusions:** Fasting plasma glucose is not meaningfully affected by prolonged walking in participants with type 2 diabetes; therefore, the reliability of fasting glucose for monitoring glycaemic burden is unlikely to be altered in patients who walk to the clinic.

**Research in context:** *What is already known about this subject?:*

- Fasting glucose is widely used to assess glycaemic control in people living with diabetes in low income countries, as HbA1c and home glucose monitoring are unaffordable. In these settings people living with diabetes will often walk long distances to receive healthcare.
- Little is known on the impact of walking on fasting glucose in people living with diabetes.

*What is the key question?:*

- Is fasting plasma glucose measure affected by a single bout of exercise such as walking in individuals with type 2 diabetes (T2D)?

*What are the new findings?:*

- There was no significant change in fasting glucose at the end of the walking exercise.
- There was no meaningful change in fasting glucose observed at any point up to 3 hours after commencing exercise.

*How might this impact on clinical practice in the foreseeable future?:*

- Fasting plasma glucose is not meaningfully affected by prolonged walking in participants with type 2 diabetes; therefore, the reliability of fasting glucose for monitoring glycaemic burden is unlikely to be altered in patients who walk to the clinic.

## INTRODUCTION

HbA1c and home capillary or subcutaneous glucose monitoring are the primary measures used to monitor glycaemic control and guide treatment titration in diabetes in high-income countries. However, these approaches are often unavailable or unaffordable [1, 2] for many of the 432.7 million people with diabetes who live in low or middle-income countries[3]. In this setting international organisations recommend the use of fasting glucose to monitor glycaemic control and titrate glucose-lowering therapy, and this remains the primary means of glucose monitoring for a substantial proportion of those living with diabetes worldwide[4].

In many low and middle-income countries, diabetes clinics are operated at regional and district hospitals, and access to transport services may be unreliable or unaffordable. Therefore, many patients will walk long distances to attend diabetes clinics[5]. While exercise such as walking result in increased glucose uptake and utilisation by the exercising muscles[6], it remains unclear whether the fasting plasma glucose measure is affected by a single bout of exercise such as walking in individuals with type 2 diabetes (T2D). We, therefore, aimed to determine the impact of walking on fasting glucose in people living with T2D.

## SUBJECTS, MATERIALS AND METHODS

### Study Design and Patients

The study was a multicentre randomised two-period crossover design to test the impact of walking on a treadmill for one hour compared to not-walking on fasting glucose changes in participants with type 2 diabetes. The Uganda Virus Research Institute (UVRI) Institutional Review Board and the Uganda National Council of Science and Technology (UNCST) approved the study, and all participants provided informed consent.

We enrolled 45 non-insulin-treated participants with T2D attending diabetes outpatient clinics at one rural and one urban hospital in Uganda. Exclusion criteria included pregnancy, acute illness, and clinical need to immediately increase their glucose-lowering medication. All participants provided written informed consent before participating.

The study was registered in the Pan African Clinical Trial Registry (https://pactr.samrc.ac.za/) (PACTR202009486614518). The overview of the study design is presented in Figure 1.

**Figure 1:**
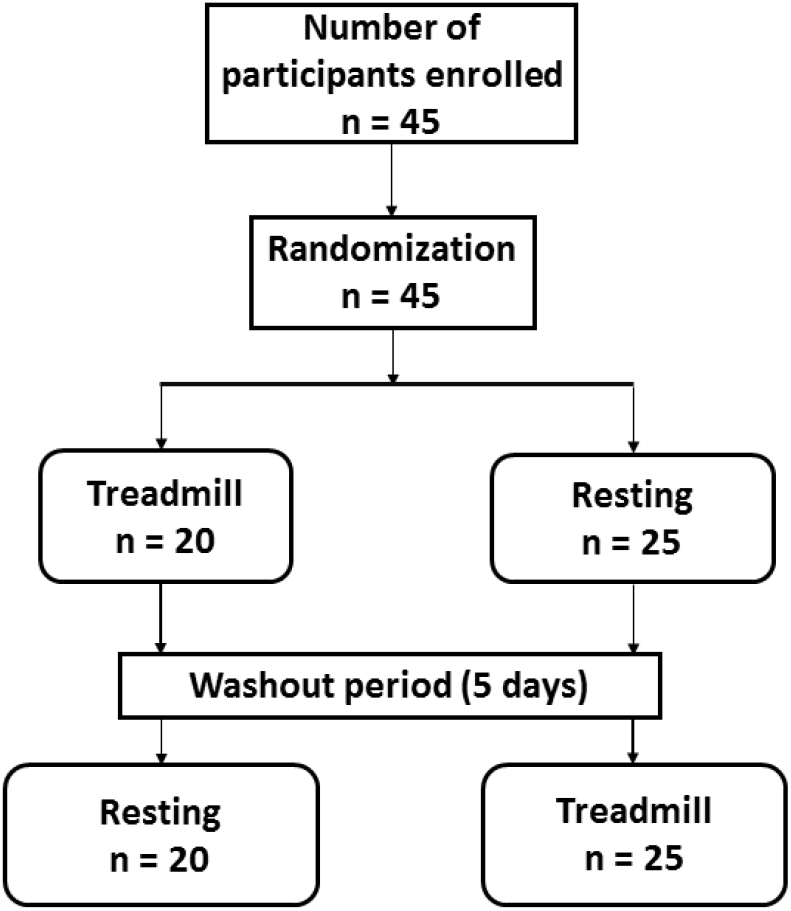
Participant flow chart. Participants attended two study visits: Treadmill visit; participant walks on a treadmill at a speed of 4.5 Km/hr for 60 minutes and then rests for the next 2 hours, resting visit; participant rests for 3 hours with minimal activity. The primary outcome was a change in fasting glucose between the walking and rest visit at 1 hour and 2-hour post treatment.

### Study visits and procedures

Participants attended two study visits: an exercise visit and a resting visit, in random order, as assigned by an independent person using computer-generated random numbers. For both visits participants attended fasting (>8 hours) before 9 am. Activity pre visit was minimised by funding visit transport and providing specific participant advice to minimise activity, and participants withheld their glucose lowering medications on the morning of the visits. There was a five-day washout period between visits. Blinding was not possible for this study.

### Exercise visit

Participants exercised at a moderate walking pace of 3 mph (4.5 km/hour) on a treadmill for 1 hour. This speed was chosen to represent average walking speed[7]. Blood samples were taken for laboratory glucose measurement at 0, 30 and 60 minutes, and post-exercise (resting) every 30 minutes for a further 2 hours (3 hours total monitoring).

### Rest visit

Participants rested (while seated) for 3 hours. Samples were taken for laboratory glucose measurement at 0, 30 and 60, 90, 120, 150 and 180 minutes.

### Primary Outcome

The pre-specified co-primary outcomes were fasting plasma glucose at 60 minutes and 120 minutes post-commencing the intervention during the walking and rest visits.

### Laboratory Analysis

Blood samples for glucose measurement were collected in a vacutainer with sodium fluoride (NaF), centrifuged and separated into two cryovials (aliquots) immediately and kept in an icebox at 4 – 8° C before being transported to the central laboratory for immediate testing (within 8 hours of collection). Plasma glucose was measured by the glucokinase method on a Roche cobas 6000 analyser, (Hitachi high technologies corporation, Tokyo, Japan) at the central Biochemistry and clinical diagnostic laboratory services (CDLS) laboratory at the MRC/UVRI & LSHTM Research Unit Entebbe Uganda.

### Statistics

The study aimed for a sample size of at least 38 with the primary outcome, which would have 80% power to detect a 1mmol/L (0.47 standard deviation) difference in fasting glucose change at 1 and 2 hours between the walking and rest visit with an alpha of 0.05.

We assessed carryover effect (occurs when the effect of the first treatment continues until the next period and alters the effect of the subsequent treatment)[8] and period effect (effect of the same treatment received at two different periods is different for each periods)[8] using mixed-effects models with 60-minute glucose as the outcome, intervention group, period, and intervention group*period interaction as independent variables, and participant ID as the random effect. Without excluding any data, we compared the difference in glucose change from baseline between resting and exercising visits, at 60 and 120 minutes from the start, using paired t-tests. The overall impact of walking (across all the time points 0-180 minutes) was assessed using mixed-effect models adjusted for baseline (time 0) glucose with time point as fixed effects and patient as random effects. We also examined whether the exercise intervention explained further variability in glucose over the total duration using the likelihood ratio test to compare with the rest intervention model.

## RESULTS

All 45 recruited participants completed both study visits (Figure 1). The characteristics of study participants are shown in table 1. There was no evidence of period effect (p = 0.29) or carryover effect (p = 0.56).

**Table 1:**
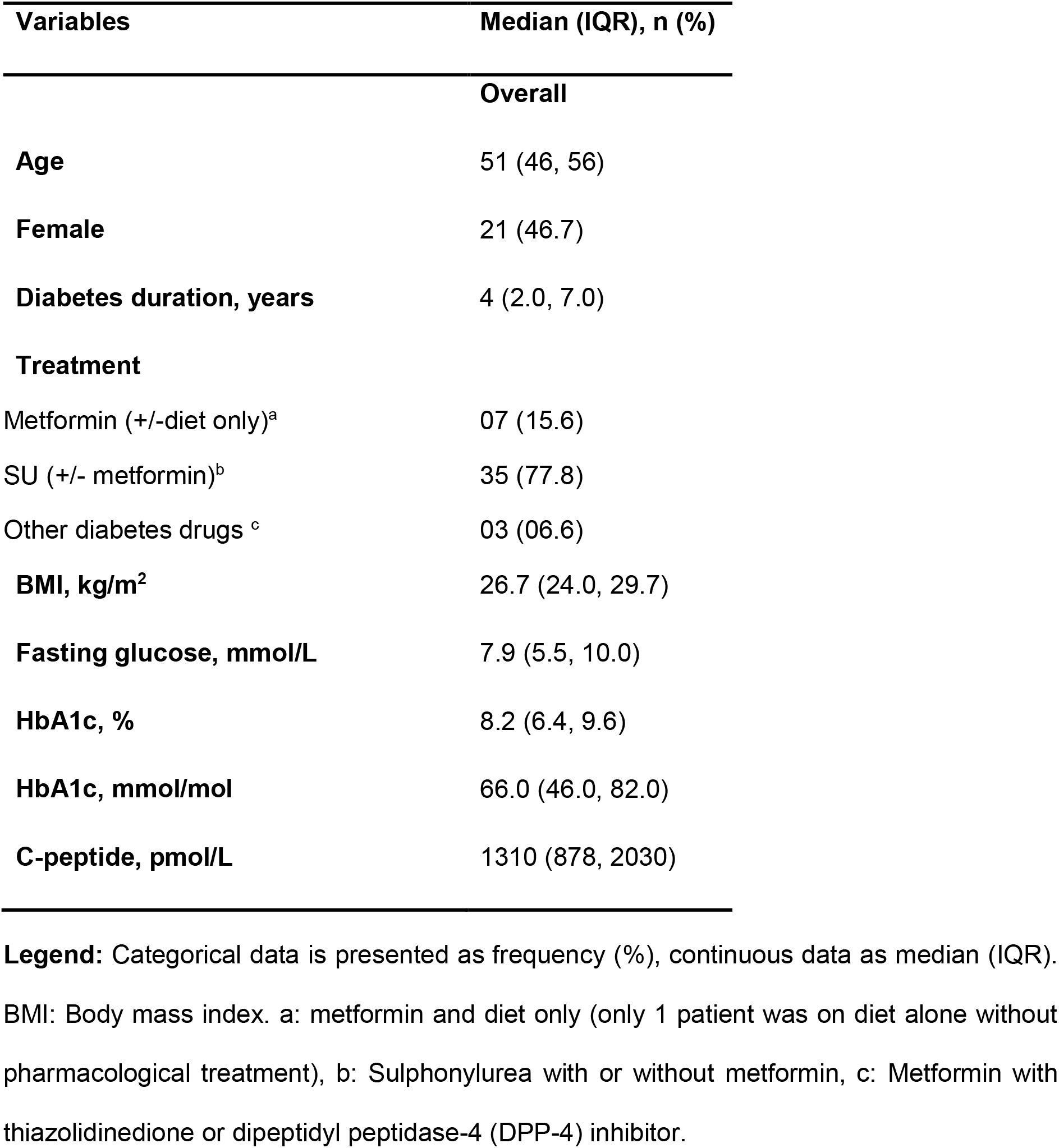
Baseline characteristics (n = 45)

### Walking was not associated with a change in fasting glucose at the end of the exercise

Walking for 1 hour was not associated with changes in fasting glucose after 60 minutes of exercise or after an additional hour of rest. Compared to the resting (control visit) glucose change from baseline (pre-intervention) with exercise was -0.15 (95% CI: -0.55, 0.26) mmol/L (p=0.48) and -0.10 (95% CI: -0.50, 0.31) mmol/L (p=0.64) at 60 and 120 minutes, respectively (Figure 2). Glucose difference was similar across all other post-baseline time points (Figure 2). The absolute values for exercise and rest visits separately are shown in figure 3.

**Figure 2:**
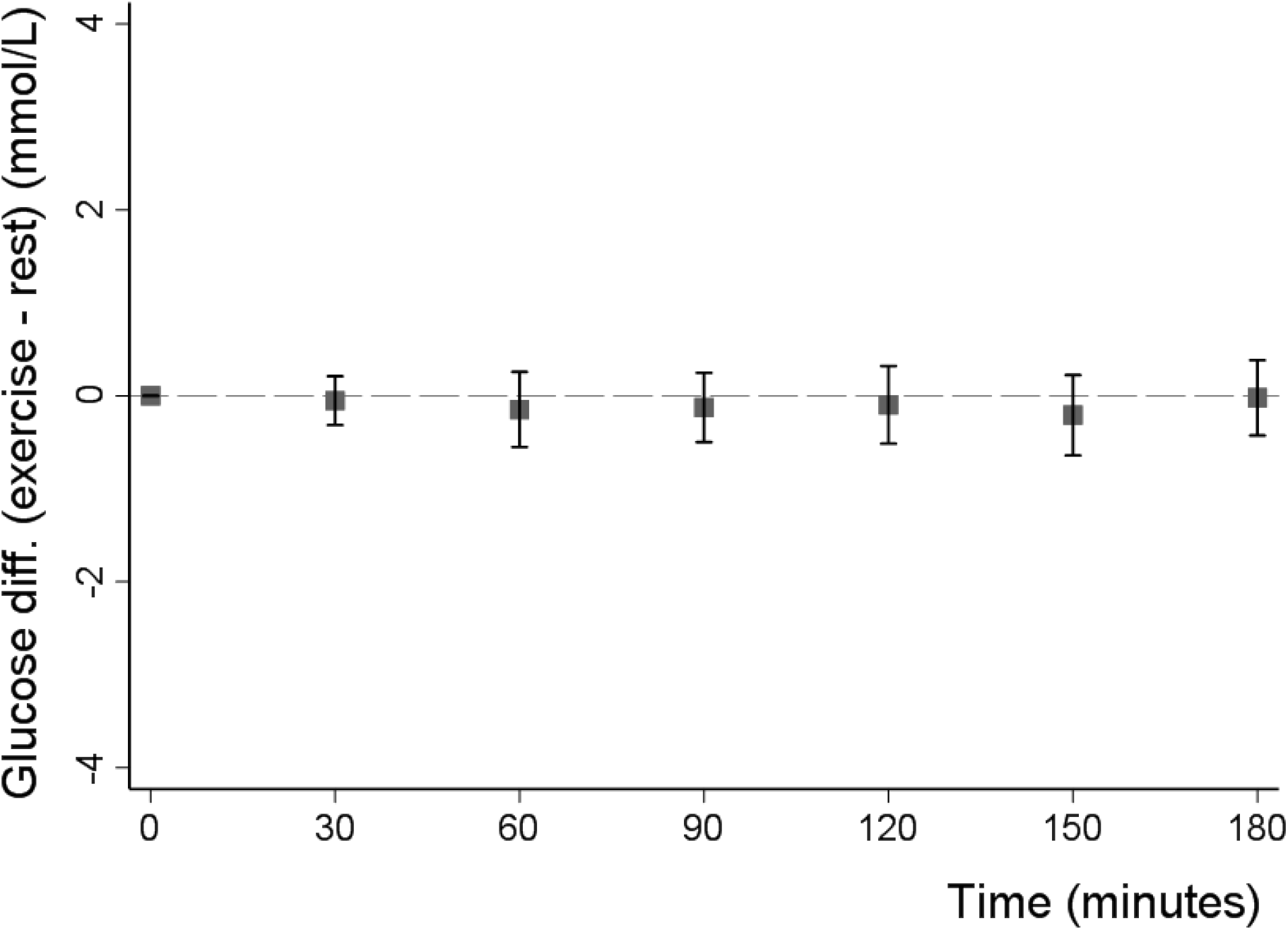
Mean difference (with 95% CIs) in glucose change from baseline between the exercise (walking) and resting visits. Y-Axis shows the difference in glucose change from baseline between the exercise and rest visits (exercise minus rest). The X-axis shows time in minutes from baseline (0 minute) up to 180 minutes from the start of the visits.

**Figure 3:**
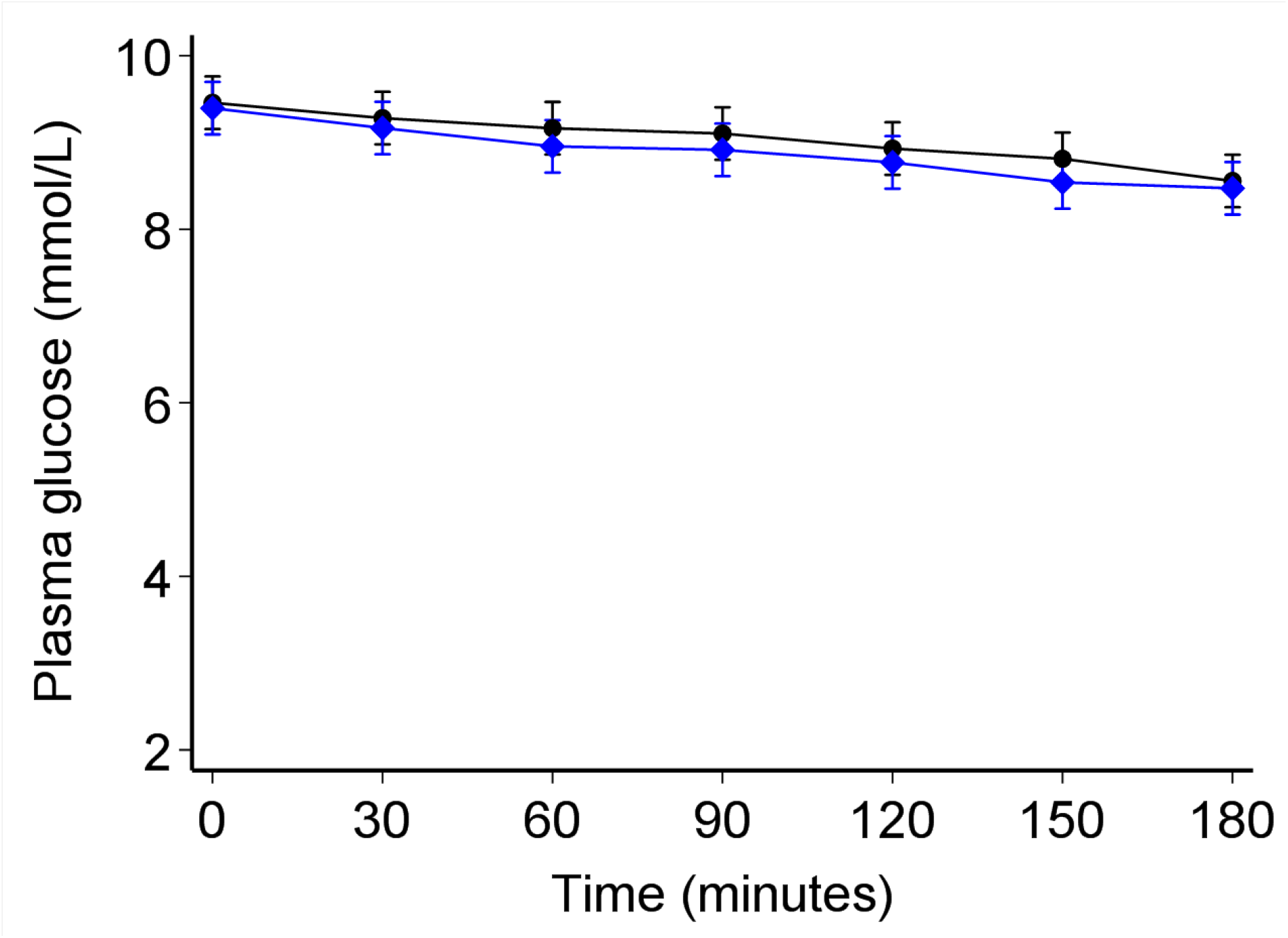
Glucose trends between the exercise (Grey diamonds) and resting visit (Black circles) after adjusting for the baseline differences between the two visits. Adjusted means adjusting for baseline differences at the two visits. The error bars denote 95% confidence intervals.

#### Walking was not associated with differences in overall post baseline glycaemia

When assessing all time points using a mixed-effects model, there was no difference in glucose between visits (p=0.67) over the 3 hours post-baseline (Figure 3). The addition of exercise into the model did not explain further variability in glucose (LR test chi2=9.0, p=0.25).

## DISCUSSION

Our findings demonstrate that in people with type 2 diabetes, fasting glucose is not meaningfully altered by 1 hour of walking.

There has been limited study of the effects of single bouts of exercise on fasting glucose in type 2 diabetes, with a recent systematic review identifying only 1 study that examined exercise in the fasting state with an appropriate comparison [9]. Karstoft et al examined the impact of continuous walking and interval walking for 1 hour (60 minutes) in a crossover study of 10 participants, and found no impact on fasting plasma glucose in comparison to the resting state [10]. Consistent with our finding, studies assessing the overall effect of exercise (of any type) on glucose measured by continuous glucose monitoring (CGM) in those living with type 2 diabetes have also shown no acute effect on fasting glucose [9, 11]. In contrast, previous studies have shown clearly that post-prandial glucose is substantially impacted by a single-bout of exercise [10, 12, 13].

Strengths of our study in comparison to previous work include; we have studied a far larger population using a randomised cross over design, and measured both pre-and post-exercise glucose and therefore we were able to control for baseline difference in the exercise and non-exercise (control) arms.

The stable fasting glucose in the walking (exercising) visit suggests that short-term moderate exercise does not affect fasting glucose, despite the known insulin independent effects of exercise on muscle glucose uptake [14]. The mechanisms underlying this finding are unclear but may involve counter-regulatory processes that favour fasting glucose stabilisation through glycogenolysis and increased mobilisation of alternative fuel sources such as free fatty acids (FFAs) from triglycerides in the adipose tissue [10, 15, 16].

These findings show that the common occurrence of patients walking long distance to clinic is unlikely to alter fasting glucose even in those with poor glycaemic control, and therefore should not affect test interpretation. They may also have implications for those undertaking moderate intensity exercise in the fasting state who are treated with sulfonylurea therapy, with recent guidance recommending carbohydrate pre-exercise in those using SUs (insulin secretagogues) regardless of fasting state [16]. The vast majority of our study participants received long acting first generation sulfonylurea treatment, but did not experience decline in glucose during exercise. However this should be interpreted with the important caveat that hypoglycaemia treatment was withheld on the morning of the study visits.

In conclusion, fasting plasma glucose is not meaningfully affected by prolonged walking in participants with type 2 diabetes; therefore, the reliability of fasting glucose for monitoring glycaemic burden is unlikely to be altered in patients who walk to the clinic.

## Data Availability

Data analysed in this study are not available for public use but to researchers upon reasonable request from the corresponding author.

## Acknowledgements

The authors are grateful to A Karungi and W Ayuru, the research assistants that assisted with data collection at the diabetes clinics. We thank the study participants and staff of the diabetes clinics of Masaka regional referral hospital and St. Francis hospital, Nsambya in Uganda.

## Contribution statement

AJN, BS, ATH, MJN and AGJ conceptualised and designed the study. AJN researched the data with assistance from BS, MJN and AGJ. AJN and LR conducted data cleaning. AJN analysed the data with assistance from LRR, RA, BS and AGJ. All authors offered advice on the study design, analysis and interpretation of the results. AJN drafted the paper which was critically revised by all authors. All authors approved the final manuscript.

## Ethics approval

The study was approved by the Uganda Virus Research Institute (UVRI) REC (UVRI-121/2019) and the Uganda National Council of Science and Technology (HS 2588).

## Funding

This work was supported by the National Institute of Health Research, UK (award reference 17/63/131). The funder was not involved in the design of the study, data collection, data analysis, and preparation of the manuscript.

## Declaration of Competing Interest

The authors declare that they have no known competing interests.

## Notes

### Competing Interest Statement

The authors have declared no competing interest.

### Clinical Trial

Pan African Clinical Trial Registry (https://pactr.samrc.ac.za/) (PACTR202009486614518)

### Author Declarations

Uganda Virus Research Institute (UVRI) REC (UVRI-121/2019) and the Uganda National Council of Science and Technology (HS 2588)

